# Comparison of Efficacy of Dexamethasone and Methylprednisolone in Improving the Partial Pressure of Arterial Oxygen and Fraction of Inspired Oxygen Ratio among COVID-19 Patients

**DOI:** 10.1101/2020.10.06.20171579

**Authors:** Muhammad Asim Rana, Mubashar Hashmi, Rizwan Pervaiz, Ahad Qayyum, Muhammad Saleem, Muhammad Faisal Munir, Muneeb Ullah Saif

## Abstract

**Introduction:** The severe acute respiratory syndrome-coronavirus-2 (SARS-CoV-2), causing coronavirus disease 2019 (COVID-19), created a pandemic in late 2019. Acute respiratory distress syndrome can occur in patients with COVID-19 due to viral replication and an uncontrolled immune reaction. Therefore, antiviral and anti-inflammatory treatments are of particular interest to clinicians. We compared the efficacy of methylprednisolone and dexamethasone in reducing inflammation and improving the partial pressure of arterial oxygen and fraction of inspired oxygen (PaO2/FiO2 or P/F) ratio in COVID-19 patients.

**Methods:** We selected 60 files for this retrospective quasi-experimental study using a convenient sampling technique and divided them into two groups of 30 patients each who had received either dexamethasone or methylprednisolone. The data were taken from the medical records of the treated patients. Group 1 patients were given dexamethasone 8 mg twice daily, and Group 2 patients were given methylprednisolone 40 mg twice daily for eight days during their stay in our high dependency unit and our Intensive Care Unit. The remaining treatment was the same for both groups using antibiotics and anticoagulation. We reviewed C-reactive protein (CRP), serum ferritin level, and P/F ratio before and after the administration of both drugs for eight days. We used a paired t-test to assess the effectiveness of both drugs on the P/F ratio of participants.

**Results:** The initial mean CRP level of Group 1 was 110.34 mg/L, which decreased to 19.45 mg/L after administration of dexamethasone; similarly, the CRP of Group 2 was 108.65 mg/L, which decreased to 43.82 mg/L after administering methylprednisolone for eight days. Both dexamethasone and methylprednisolone significantly improved the P/F ratio (p<0.05), and dexamethasone was significantly more effective than methylprednisolone (p<0.05).

**Conclusion:** Steroids’ ability to reduce inflammation and suppress the immune response make them an effective tool in the treatment of COVID-19. Steroid therapy is effective in controlling inflammation markers, and, specifically, dexamethasone is effective in improving the P/F ratio in COVID-19 patients. Physicians should consider the use of dexamethasone use in appropriate patients with COVID-19.

## Introduction

The coronavirus disease-2019 (COVID-19) pandemic caused by the severe acute respiratory syndrome-coronavirus-2 (SARS-CoV-2) originated in Wuhan, China, in December 2019. Although most infected patients undergo an uneventful recovery, approximately 19% of patients experience severe pneumonia and 14% experience a progressive worsening to critical pneumonia [1].

Patients with severe COVID-19 quickly progressed to acute respiratory failure, pulmonary edema, and acute respiratory distress syndrome (ARDS) [2], which occurred not just because of an uncontrolled viral replication but also because of an uncontrolled immune reaction from the host. With the existence of uncontrolled viral replication, the presence of an increased number of damaged epithelial cells and cell debris activate a massive cytokine release, also known as a cytokine storm, with hyperinflammation and immune inhibition, characterized by decreased memory cluster of differentiation-4 þ T helper cells and an increased cluster of differentiation-8 cytotoxic activity [3].

As a result, antiviral and anti-inflammatory treatments have become an increasing concern for clinicians [4]. A randomized clinical study demonstrated that corticosteroid therapy could reduce inflammatory responses, reduce treatment failure, and reduce the time to clinical stability in community-acquired pneumonia without major adverse effects [5].

Recently, Villar et al. reported that early administration of dexamethasone shortened mechanical ventilation time and overall mortality for patients with moderate-to-severe ARDS [6]. Corticosteroid therapy was associated with improved clinical outcomes in severe COVID-19 patients in clinical practice. Zhou et al. reported that corticosteroid therapy improved the clinical symptoms and oxygenation of patients with COVID-19 [7].

Wang et al. also found that, for patients with severe COVID-19, corticosteroid therapy reduced hospital length of stay and intensive care unit (ICU) stays [8]. Chinese experts considered it prudent to administer short courses of corticosteroids at low-to-moderate doses for critically ill patients with COVID-19 [9]. Wu et al. reported that treatment with methylprednisolone decreased the risk of death for individuals with COVID-19 with ARDS [10].

### Objective

Using steroids in moderate to severe disease is a standard practice in management of COVID19 and we have been following the same. Some cases were treated with methylprednisolone where others were treated with equivalent doses of dexamethasone. This has automatically created a patient pool consisting of two groups.

The objective of this study was to assess the effectiveness of dexamethasone and methylprednisolone in COVID-19 patients as well as compare both drugs in regards to partial pressure arterial oxygen and fraction of inspired oxygen (PaO_2_/FiO_2_ [P/F]) ratio improvement. A significant amount of research has been published on the effectiveness of steroids in COVID-19 patients, but there is limited literature on comparing the effectiveness of dexamethasone and methylprednisolone.

### Hypothesis

We created two hypotheses: the H1 (alternate hypothesis) and the H0 (null hypothesis). The H1 hypothesis states that methylprednisolone and dexamethasone can improve the P/F ratio in COVID-19 patients. The H0 hypothesis states that methylprednisolone and dexamethasone cannot improve the P/F ratio in COVID-19 patients.

## Materials and Methods

We used a retrospective quasi-experimental study design to assess the effectiveness of methylprednisolone and dexamethasone in the improvement of the P/F ratio in COVID-19 patients. We used a convenient sampling technique to select files (medical record) of 60 participants, all of whom had been admitted and treated in the HDU/ICU and had been on bi-level positive airway pressure. These 60 patients (medical records) were divided into two groups: Group 1 who had received dexamethasone, and Group 2 who had received methylprednisolone. Each group consisted of 30 participants (medical records of 30 patients). It needs to be reemphasized that these cases were already treated and had been concluded. They were treated by different treating teams. One group was pro-dexamethasone who used to give dexamethasone to their patients while other team had been favoring and used methylprednisolone in equivalent doses to dexamethasone. Apart from the type of steroid used all other management was same for both groups of patients including anticoagulation and we would like to declare that both groups were on therapeutic anticoagulants. We had administered dexamethasone 8 mg twice daily to one Group (labelled as Group 1) participants and methylprednisolone 40 mg twice daily (almost equivalent dose to dexamethasone) to now labelled as Group 2 participants. The duration of treatment compared was for eight days. Laboratory specimens were being sent daily during the morning shifts to enable the treating clinicians to take decisions during their treatment periods. We have chosen two sets laboratory values for this study one set which was taken on the first day of hospitalization and second set from eighth day of administration of drugs, from their medical records. Similarly for the P/F ratio comparison, we took the PaO_2_ values from their daily morning arterial blood gas reports, two sets again. For data analysis, we described demographic data in descriptive form (frequencies and percentages); laboratory values were analyzed in the form of mean, and comparison between dexamethasone and methylprednisolone effectiveness of the P/F ratio was analyzed using a paired t-test.

## Results

Our results are presented in three parts. Part I includes demographic characteristics of participants, Part II includes outcomes of laboratory value changes in response to dexamethasone and methylprednisolone, and Part III includes a comparison of outcomes in response to both drugs.

### Part I

Sixty patients’ medical records was selected for this study, with 30 participants in each group. Group 1 which had received dexamethasone for eight days, and Group 2 which had received methylprednisolone for eight days. Both groups were on therapeutic anticoagulants. In Group 1, 33.33% were women, and 66.67% were men; similarly, in the Group 2, 30% were women, and 70% were men (Figure 1). The mean age of Group 1 was 53.8 years, and the mean age of Group 2 was 53.9 years (Table 1).

**Table 1.** Mean age.

|  | Mean Age of Group 2 | Mean Age of Group 1 |
| --- | --- | --- |
| Mean | 53.8 | 52.9 |

**Figure 1.**
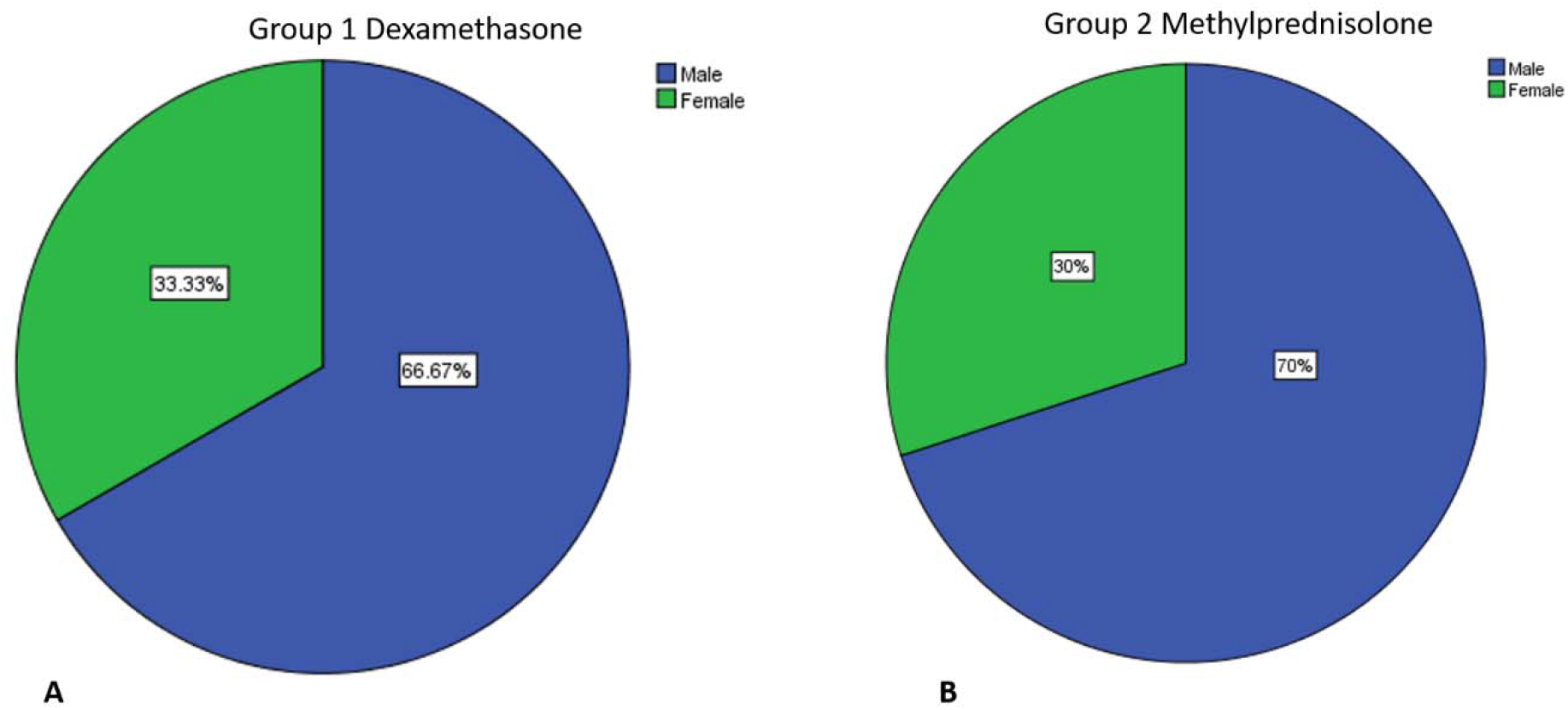
Sex distribution for Group 1 (A) and Group 2 (B)

### Part II

In this section, we discussed the changes that occurred in laboratory values of patients before and after administration of dexamethasone and methylprednisolone and how effectively dexamethasone and methylprednisolone worked. In Table 2, the CRP difference is shown. The initial mean CRP of Group 1 was 110.34 mg/L, which decreased to 19.45 mg/L after administration of dexamethasone and was considered a good reduction in CRP value. Mean inflammation was reduced in the maximum range; similarly, the CRP of Group 2 was 108.65 mg/L, which decreased to 43.82 after the administration of methylprednisolone for eight days. In Table 3, serum ferritin levels are shown. The initial mean serum ferritin of Group 1 was 763 ng/mL, which decreased to 494.30 ng/mL after administration of dexamethasone for eight days. Similarly, the serum ferritin level of Group 2 was 631.53 ng/mL, which decreased to 493.07 ng/mL after administration of methylprednisolone for eight days.

**Table 2.** C-reactive protein before and after steroid therapy.

|  | Mean | N |
| --- | --- | --- |
| CRP before dexamethasone | 110.3467 mg/L | 30 |
| CRP after dexamethasone | 19.4573 mg/L | 30 |
| CRP before methylprednisolone | 108.6543 mg/L | 30 |
| CRP after methylprednisolone | 43.8297 mg/L | 30 |

Abbreviations: CRP, C-reactive protein.

**Table 3.** Ferritin levels before and after steroid therapy.

|  | Mean | N |
| --- | --- | --- |
| Ferritin pre-dexamethasone | 763.00 ng/mL | 30 |
| Ferritin post-dexamethasone | 494.30 ng/mL | 30 |
| Ferritin pre-methylprednisolone | 631.53 ng/mL | 30 |
| Ferritin post-methylprednisolone | 493.07 ng/mL | 30 |

The focus of this study was the evaluation of the P/F ratio in both groups after the administration of methylprednisolone and dexamethasone. According to Table 4, the P/F ratio of Group 1 at the initial stage was 118.20, improved to 170.41 after the administration of dexamethasone for eight days. Both values were in the range of moderate respiratory failure, but the P/F ratio had improved. Similarly, in Group 2, the P/F ratio at the initial stage before methylprednisolone therapy was 105.66 and improved to 136.25 after the administration of methylprednisolone.

**Table 4.** P/F ratio before and after steroid therapy.

|  | Mean | N |
| --- | --- | --- |
| P/F ratio pre-dexamethasone | 118.2067 | 30 |
| P/F ratio post-dexamethasone | 170.4130 | 30 |
| P/F ratio pre-methylprednisolone | 105.6609 | 30 |
| P/F ratio post-methylprednisolone | 136.2575 | 30 |

Abbreviations: P/F, partial pressure arterial oxygen and fraction of inspired oxygen ( $\text{PaO}_2/\text{FiO}_2$ ).

### Part III

This section consisted of outcomes comparison after administering methylprednisolone and dexamethasone. As mentioned previously, the main focus of this study was to assess the effectiveness of dexamethasone and methylprednisolone on the P/F ratio. To compare the pre- and post-P/F ratio, we applied a paired sample t-test to check the effectiveness of both drugs. Findings revealed that the mean P/F ratio after dexamethasone (170.4130) was significantly higher than the mean P/F ratio before dexamethasone (118.2067; p=.000). Similarly, the P/F ratio after methylprednisolone (136.2575) was higher than the P/F ratio before administration methylprednisolone (105.6609) for eight days (p=.009; Table 5). As these p-values indicate, dexamethasone improved the P/F ratio significantly more than methylprednisolone.

**Table 5.**
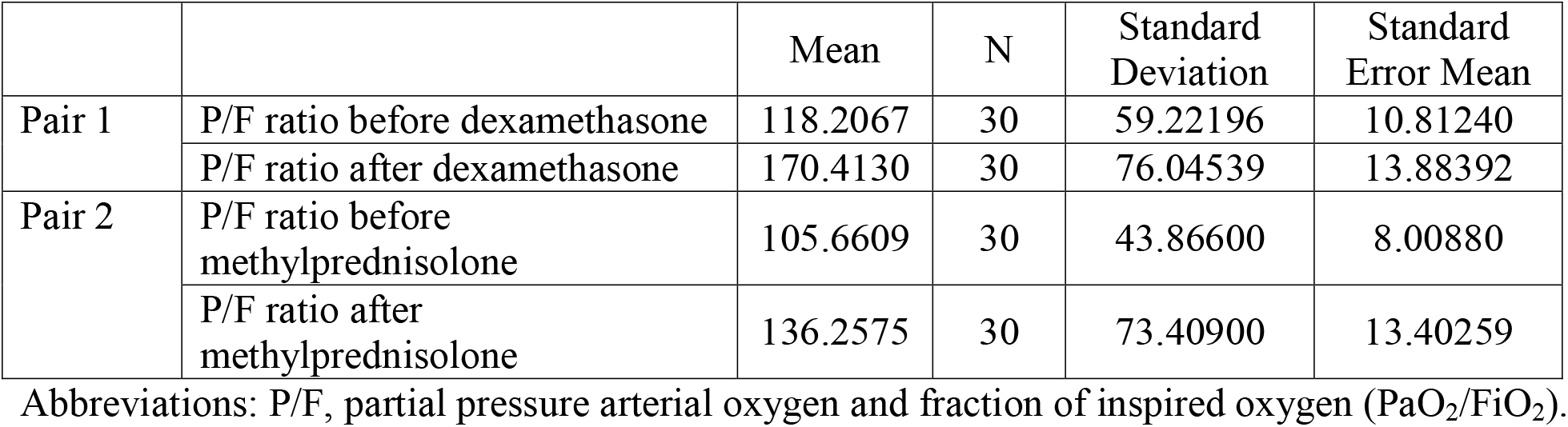
Paired sample statistics.

**Table 6.** Paired samples test.

|  |  | Paired Differences |  |  |  |  | t | df | Sig.<br>(2-tailed) |
| --- | --- | --- | --- | --- | --- | --- | --- | --- | --- |
|  |  | Mean | Standard<br>Deviation | Standard<br>Error<br>Mean | 95%<br>Confidence<br>Interval of the<br>Difference |  |  |  |  |
|  |  |  |  |  | Lower | Upper |  |  |  |
| Pair 1 | Pre-P/F ratio<br>dexamethasone -<br>post-P/F ratio<br>dexamethasone | -<br>52.20 | 50.070 | 9.141 | -<br>70.902 | -<br>33.509 | -<br>5.711 | 29 | .000 |
| Pair 2 | Pre-P/F ratio<br>methylprednisolone<br>post-P/F ratio<br>methylprednisolone | -<br>30.59 | 60.268 | 11.003 | -<br>53.101 | -8.091 | -<br>2.781 | 29 | .009 |

Abbreviations: t, difference relative to the variation in data; df, degrees of freedom; P/F, partial pressure arterial oxygen and fraction of inspired oxygen( $\text{PaO}_2/\text{FiO}_2$ ).

The H1 alternate hypothesis (methylprednisolone and dexamethasone are effective in improving P/F ratio in COVID-19 patients) can be accepted, and the H0 null hypothesis (methylprednisolone and dexamethasone could not improve the P/F ratio in COVID-19 patients) can be rejected on the basis of the paired t-test.

## Discussion

The mean age of patients in this study was 53.3 years, and most patients (66.67%) were men, and 33.33% were women. Spagnuolo et al. showed that the median age of participants was 63.5 years (53.5 to 74.0 years), and 34% of patients were older than 70 years [11].

We found that corticosteroids reduced CRP levels. A similar study conducted in 2005 indicated that the withdrawal of inhaled corticosteroids increased serum CRP levels. The reintroduction of the inhaled steroids suppressed the CRP levels. Hence, corticosteroids can reduce serum CRP and other circulating inflammatory cytokine levels in some acute inflammatory states [12].

In the present study, the initial mean serum ferritin in both drug groups were reduced after eight days. A study conducted on *Mycoplasma pneumoniae* in November 2015 elaborated that ferritin levels may be useful as indicators of the severity of pneumonia for initiation of corticosteroid therapy [13].

The findings of the present study revealed that both dexamethasone and methylprednisolone were effective in improving the P/F ratio. Spagnuolo et al. assessed the effectiveness of steroids in COVID-19 patients, where differences between patients taking steroids and patients not taking steroids were observed [11]. A remarkable improvement in P/F ratio was observed. The proportion of patients with a baseline PaO_2_/FiO_2_ ≤ 200 mmHg was 45.8% for patients taking steroids compared to 34.4% in patients not taking steroids (p=0.023). The proportion of patients with a baseline PaO_2_/FiO_2_ ≤100 mmHg was 16.9% for patients taking steroids compared to 12.7% who were not taking steroids (p=0.027). Steroid use also decreased the length of hospitalization (20 vs. 14 days; p<0.001) [11].

There were certain limitations of the present study. We did not include a control group, did not assess any underlying comorbidities, and did not compare the effectiveness of steroid therapy in regard to age, sex, and severity of illness. Randomized controlled clinical trials are required to confirm the effectiveness and safety of corticosteroid therapy and to further study the long-term outcomes after discharge with a large sample size for better generalization of findings.

## Conclusion

Steroids have long been known to reduce inflammation and suppress immune reactions. These actions make steroids an effective tool in the treatment of COVID-19, and they have become a standard of treatment all over the world. Dexamethasone is more effective in improving the P/F ratio in COVID-19 patients compared to methylprednisolone. Physicians should consider the use of dexamethasone use in appropriate patients with COVID-19.

## Data Availability

All data is available with the IRBEC of hospital.

